# *My family sold a cow to pay for my Traditional doctor and now there’s no money to travel to the HIV clinic*: barriers to antiretroviral adherence among rural-Indigenous peoples living with HIV in the Comarca Ngäbe-Buglé, Panamá

**DOI:** 10.1101/2021.12.01.21267149

**Authors:** Amanda Gabster, Eliana Socha, Juan Miguel Pascale, Gonzalo Cabezas Talavero, Alezander Castrellón, Yaremis Quiel, César Gantes, Philippe Mayaud

## Abstract

**Introduction:** The Comarca Ngäbe-Buglé (CNB) is an administratively autonomous Indigenous region in Western Panama that is home to over 200,000 individuals of Ngäbe and Buglé ethnicities. The CNB is the most impoverished region in Panama and is relatively isolated from outside influences, with limited roads, electricity, and internet connection. Around 1.5% of all rapid HIV tests are positive, compared to a national prevalence of 0.9%; in CNB, diagnosis tends to be late where 56.3% of individuals had an initial CD4 count of <350 cells/mm^3^. In this region, antiretroviral treatment (ART) dropout is five times higher than the national average; there is high early mortality due to opportunistic infections. This study aims to describe some of the barriers associated with ART adherence and retention in HIV care among PLHIV the CNB. A better understanding of factors that obstruct adherence could lead to more effective HIV care and prevention in CNB.

**Methods:** We conducted 21 semi-structured interviews with PLHIV who reside across all three regions of the CNB and who have attended an ART clinic at least once. The interviews took place between November 2018 and December 2019.

**Discussion:** Psychological health and social support and discrimination acted as both individual-level facilitators and barriers to adherence and retention. Notably, structural barriers included difficult access to ART care due to travel costs, ART shortages, and uncooperative Western/Traditional medical systems. Recommended interventions used in other Low- and Middle-Income settings include increasing peer and family-level support and community knowledge and understanding of HIV infection. Additionally, we suggest structural interventions, including decreasing cost and distance of travel to the ART clinic through decentralization of services and multi-month dispensing, decreasing food scarcity, and increasing collaboration between Western and Traditional providers.

## Introduction

The Comarca Ngäbe-Buglé (CNB) is a politically semi-autonomous region in Western Panama, home to over 200,000 people of Ngäbe and Buglé ethnicities, making up 5.4% of the total Panamanian population (1). This Comarca has the highest levels of poverty (defined as <$1.90/day) and extreme poverty (<$1.25/day) in Panama: in 2015, it was estimated that between 56 and 78% of households across the CNB lived in extreme poverty(2). In addition, the CNB is relatively isolated from the rest of Panama, with limited roads, electricity, and internet access (3).

In 2018, the HIV prevalence in Panama among 15-49-year-old individuals was 0.9%, whilst in the CNB, 1.5% of tests performed were positive (4). Since 2000, ART has been free to all HIV-positive individuals in Panama (5). Retention is the ongoing participation in HIV care, while adherence, use of ART regularly as a health professional indicates. Together, retention and adherence lead to HIV viral load suppression, reduction of ART resistance, decrease in ongoing HIV transmission (6), and overall survival (7, 8). In the CNB, access to ART and HIV-related care is only available at two of 14 health centers.

This lack of adherence and retention in HIV care may be reflected in epidemiological reports. For example, nationwide in 2018, HIV was the ninth most common cause of death among men and women, while in CNB, HIV was the first cause of death among men and second among women (9). In addition, adherence to ART has been less than optimal in the CNB. In 2019, late diagnosis for HIV was common, where 56.3% (237/421) of all patients had an initial CD4 count at <350 cells/mm^3^ and 24.7% (104/421) at <200 cells/mm^3^. That same year, 30.3% (226/747) of patients in CNB abandoned ART and HIV care--the majority within the first year after their diagnosis--compared to the national average of 6.5% (1050/16,043) (10). Furthermore, in 2019, viral suppression (<1000 copies/ml) among all patients was reported among only 52.3% (312/589) of men and 49.0% (73/149) of women. Opportunistic infections are also relatively common; 17.2% (22/128) of individuals diagnosed with tuberculosis tested positive for HIV, and 54 patients who attended the ART clinic presented with an opportunistic infection (24.1% [13/54] and diarrheal disease, 7.4% [4/54] with oral candidiasis, 7.4% [4/54] with histoplasmosis, cerebral toxoplasmosis or HIV-associated pneumonia (10). In 2018, ART shortages occurred for 1-3 months throughout the year in: efavirenz/emtricitabine/tenofovir, Lopinavir/Ritonavir (200/50mg and 80/20mg), and Raltegravir (400mg).

The Social-Ecological Theory for Health has been used previously to describe factors associated with ART adherence (11, 12). Three primary factor levels have been associated with the model: individual, social, and structural levels (13, 14). Individual-level factors include personal attitudes and beliefs; the social-level factors include interpersonal interaction with peers, friends, family, and peers; the structural-level factors include the healthcare system, poverty community organizations, and policy.

Considering the high prevalence and proportion of deaths due to AIDS-related illnesses in CNB, we undertook a qualitative study among HIV-positive individuals to describe the individual, social, structural levels facilitators and barriers to ART adherence and retention in care in CNB. A better understanding of these factors will help in the development of more effective, culturally congruent HIV care for CNB PLHIV.

## Methods

### Study design and participants

The study design and topics arose during previous research of sexually transmitted infections (STI), including HIV among young people in CNB (15). We conducted interviews among people living with HIV (PLHIV) at the two existing ART clinics in CNB or a private location of their choice in the interviewee’s community. Clinicians who provide ART (authors CG and YQ) selected participants from their clinics using purposive sampling based on varying levels of adherence and retention success obtained from clinic charts. The interview schedule **(Table 1)** was piloted among ten non-PLHIV in two CNB communities, non-PLHIV, for language understanding and acceptability before undertaking the study.

**Table 1:** Interview schedule.

| **People living with HIV** |  |
| --- | --- |
| **Key questions and themes** | **Prompts and sub-questions** |
| Can you tell me a bit about your diagnosis? What happened after? | • What was it that motivated you to get treatment?<br>• How were you treated in the health centre or hospital? |
| Tell me about the treatment you are taking now. | • Why did you select this treatment? ¿Por qué elegiste este tipo de tratamiento? Or why did you stop taking treatment or go to the ART clinic?<br>• How do you feel about taking this treatment?<br>• Have you taken *botánica*? Can you tell me about that? |
| Tell me a bit about the difficulties you have faced while trying to access ART. | • How much does it cost for you to get your ART?<br>• Have you ever had a bad experience in the system? Did this affect your ART use? |
| What has been the process of adaptation to your diagnosis | • How do you feel about your diagnosis?<br>• What plans do you have for your future? |
| How has your diagnosis changed or affected your relationship with your family or friends? | • What have been the reactions of your family and friends?<br>• How do they support you?<br>• How do they treat you differently? |
| And the rest of your community? | • Have you felt you were discriminated against? Can you tell me about it?<br>• What are the opinions of your community regarding HIV, and how do these opinions affect you? |
| What are some things that have affected your wellbeing since you were diagnosed? | • Is there something that has affected you negatively? What happened, and how did it affect you?<br>• Tell me about your spiritual beliefs and how these affect your diagnosis. |
| Tell me what you have learned about HIV since you | • What would you like to learn more about? |
| were diagnosed. | What can the clinic do better?<br>• What kind of orientation do you wish your family received? |

Authors AG, ES, AM conducted the semi-structured interviews in Spanish, which is spoken at a basic level by the majority of people in CNB (16). Interviews took 20-40 minutes and were conducted until saturation of key themes regarding ART adherence. A digital recorder was used to record the interviews. Interview themes and questions are found in **Table 2**.

**Table 2.**
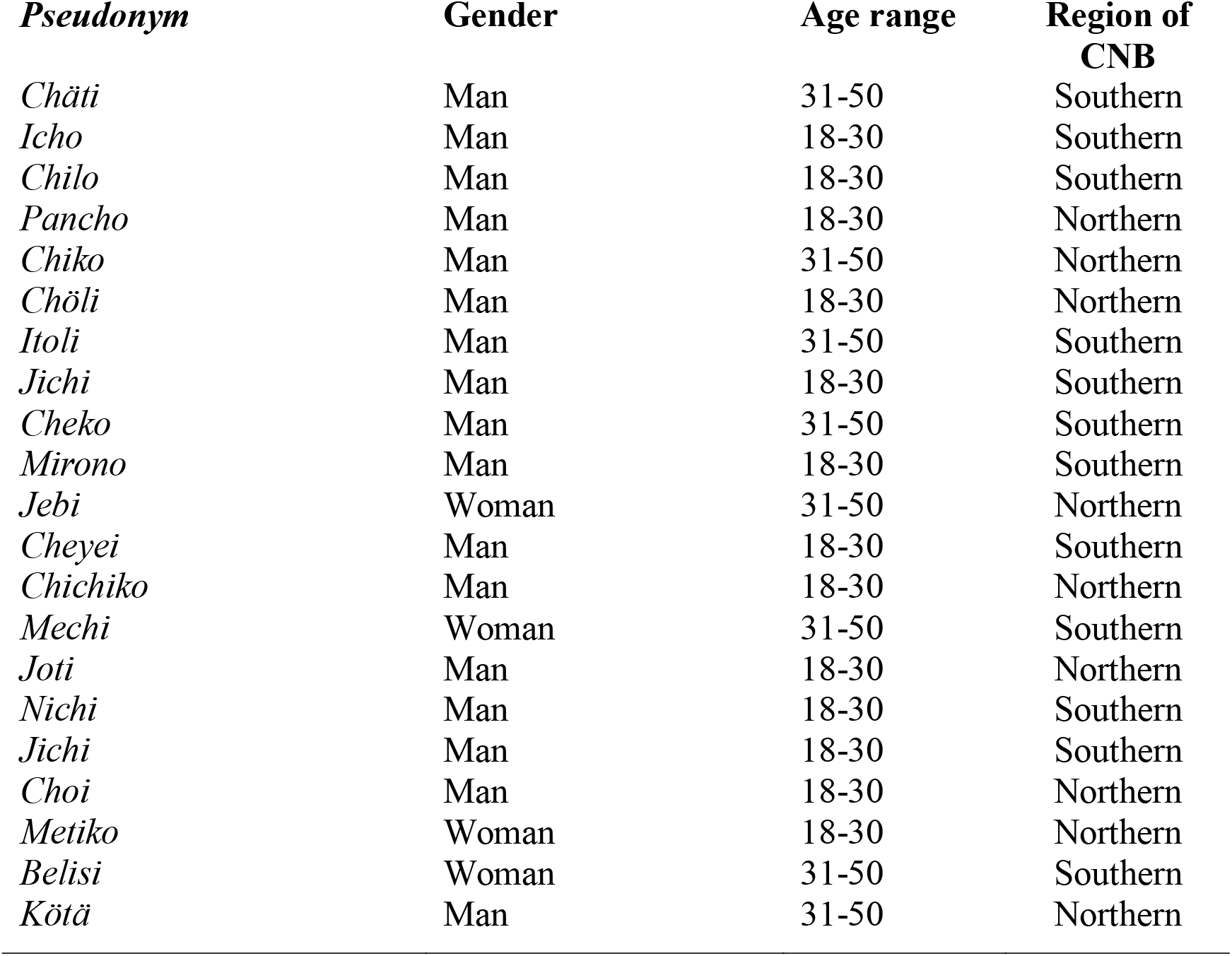
Participant charactersitcs, Comarca Ngäbe-Buglé, Panama, 2018.

### Analysis

The interviews were transcribed by authors GCT and AC and then translated from Spanish to English by AG. A research assistant cross-checked the translation. Deductive thematic analysis was used to uncover themes of individual, social and structural factors related to ART adherence. Interview transcripts were first read, then data were broken into codes using NVIVO12 software (QSR International, Melbourne, Australia). Codes were organised into themes that were part of the original codebook; other themes were allowed to emerge. AG coded interviews; four interviews were checked for interrater reliability with author ES. Contradictions were agreed upon. Saturation of themes and codes was reached within 12 interviews; coding was completed for all interviews.

### Ethical approval

The research was approved by the Comité Nacional de Bioética de la Investigación de Panamá (EC-CNBI-20180836). All participants were ≥18 years and signed an informed consent prior to initiating the interviews.

## Results

Between April and December 2019, 21 semi-structured interviews were conducted with PLHIV, 12 from the Southern mountainous regions of Nedrini and Kädriri, nine from the Northern region of Ñö Kribo. All participants were of Ngäbe or Buglé ethnicities, 81% were male (**Table 2)**; 13 (62%) had at least two missed clinic visits over the past two years for treatment collection or CD4/viral load testing.

Based on the Social-Ecological model, we discuss the facilitators and barriers to ART adherence and retention in care at the individual, social and structural levels.

### Individual and social facilitators

We found individual factors related to ART adherence included three themes: knowing how ART controls the virus, the inclusion of ART in scheduled routines, and good psychological health, which in turn seemed to be influenced by social support received from two social groups: clinic peers, and family.

Chäti, Chiko, Jichi, Icho, Belisi, Metiko, Mirono, and Jebi mentioned they knew how ART controls the infection but does not cure HIV. Belisi and Jichi similarly stated, “I know there is no cure; my medicine is to control the virus.” In addition, some participants indicated adherence was easier when including their dose as part of their daily routine. For example, Mirono, Chäti, and Metiko and Belisi indicated it was easy for them to remember to take ART because they take their dose every night before bed at the same time.

Good psychological health was found to be directly influenced by social support the participant received from clinic peers and family. For example, Mirono, Cheyei said they found social support and inspiration to maintain adherence and retention in care while talking to other patients at the clinic. In previous years, a social worker organized a temporary peer support group at the southern ART clinic. Cheko, Jichi, Chilo reported the group helped them feel supported, which, they said, translated into better retention and adherence.

Patient social support also came from family members. For example, Chiko, Chichiko, Jebi, Nechi, Mirono, Choi, Kötä, Chöli, and Itoli indicated that family support was crucial for ART adherence. Often, one family member knew of the diagnosis and supported the patient emotionally by checking in; consequently, the individual wanted to continue with ART. For Mirono, Kötä, Chiko and Jebi, family support also impacted retention in care, as family members often gave the patient financial assistance for monthly travel to the ART clinic.

### Structural facilitators

Two main structural facilitators were elicited: ART clinic expansion and nutritional supplements. The Ministry of Health provided structural support. The first ART clinic opened in 2010 in the Southern regions Nedrini and Kädriri; in 2017, a second clinic was opened in the Northern region of Ñö Kribo. The new clinics have aided in decreasing travel costs and distance for many patients. Participants also indicated that both clinics supply nutrient enhanced porridge (*crema*) to all patients at ART retrieval. Some patients (Pancho, Jichi, Belisi, and Itoli) notably said that they would not have food at night when they take their dose without the crema.

### Individual and social barriers

We found individual barriers to include lifestyle choices and psychological health. Lifestyle choices of an individual such as drinking and partying were found to increase the chances of unprotected sex (Pancho) and decrease regular adherence (Chöli, Cheyei, Pancho). Chöli said, “When there’s a party, and I drink…sometimes I forget to take my medicine.’ Despite some mentioning these lifestyle choices, most participants indicated they do not consume alcohol or party.

We found evidence that individual factors, such as the patient’s psychological health, influenced feelings of depression and loss of motivation for adherence. In addition, psychological health was founded in discrimination from various aspects of the participant’s social circle: family, friends, community members, and religious groups, which in turn impacted the patient’s ART retention and adherence.

Family-level discrimination was felt by Chiko, Choi, Cheko, Nichi, Pancho, Jichi, and Itoli. These participants indicated a link between family discrimination and feelings of isolation and depression, which in turn led to apathy towards ART adherence and retention. Chiko, Choi, Cheko, Nichi, Pancho, and Jichi told of being segregated from their family during day-to-day routines, including while cooking, sharing of food, and sleeping spaces. Cheko explained the effects of family-level discrimination on ART adherence: “Your family is your base, if your family rejects you, what kind of strength do you have to live?… then you don’t take your medicine and you get an infection and die.”

Other patients feared community status disclosure and community-level discrimination. Mechi, Cheyei and Mirono said they worried community members would raise questions about their whereabouts when they left their community every month on the same date. Mirono, Jebi, Cheyei, Mechi, Joti, Kötä, Metiko, Cheko, Jichi, and Choi said they had felt discrimination from friends and community members. Many indicated this led them to isolate, and not seek further social interaction. Participants, Joti, Icho, Belisi, Jichi, and Choi reported that despite community-level discrimination, the emotional support they received from their family counteracted feelings of isolation; adherence and retention were not affected.

### Structural barriers

Structural barriers to ART adherence included four general themes: friction between Western/Traditional systems, food security, medication shortages, difficulties of access due to distance and cost.

Many participants felt they had to choose between Western and Traditional medicine for four reasons: 1) both Traditional and Western providers told them not to use pluralistic treatments simultaneously. For example, Poncho and Kötä indicated Traditional providers told them not to mix treatments because ART would lessen the effects of *botanica* (herbal medicine); Chichiko, Itoli, Chilo, Jebi, Belisi, and Cheko said Western doctors feared unknown side-effects from mixing treatments. 2) Due to the high price of some Traditional providers, Joti, Cheyei, and Jichi were left without traveling money to go to the ART clinic. For example, Joti said, ‘my family wanted me to take *botanica*, they sold a cow to pay for it, but then we didn’t have more money to travel to the clinic’. 3) Jebi, Nichi, Chilo Pancho, and Chöli reported that Traditional providers offered a cure for HIV. Chilo and Chöli even indicated they had stopped treatment at some time because they believed they could be cured. Participants reported that some Traditional providers told them they could be cured by believing that God and *botanica* would heal them.

A second structural barrier for several patients was food security. Icho, Pancho, Chichiko, Choi, and Kötä said they were told to take their pills with food, but often they did not eat in the evenings. Chichiko notably said, “They told me to eat before I take my pills, but my family doesn’t eat at night… So I didn’t know if I should take my medicine.” To lower side-effects of ART, patients are advised to take medicine at night with food, however traditionally, and perhaps due to poverty, many individuals in CNB eat only one meal a day.

All participants interviewed spoke of having their treatment interrupted at some time due to supply shortage at the ART clinic they attend. Notably, Cheko said, “In 16 years, I’ve been on treatment, almost yearly the clinic runs out of pills for a while. Sometimes I don’t get my pills for a month or three, sometimes they say to come the following week to pick them up, but then I don’t have money to travel again… when the pills come in and I can travel, I start taking them again.”

The last structural barrier we found is access to the ART clinic due to the distance and the cost of travel. Kötä, for example, explained he walks 8 hours to get to a road where there is a bus down to the clinic. Metiko and Chöli indicated that because of the distance, they could not travel to and from the clinic in one day. Therefore, further costs are incurred with finding where to stay, or more disconcertingly, they may sleep outside the clinic (Pancho). Chilo, Chäti and Jichi said they rely on family for money to travel; however, at times, family members cannot provide for the transport costs; therefore, the patient may miss picking up the treatment that month, or altogether drop out of care. Chäti notably said, “sometimes my father tells me it’s too expensive (to keep me in care), so I want to stop taking pills to not be a burden.”

## Discussion

The Comarca Ngäbe-Buglé (CNB) is an isolated region of Panama, with poor access to ART and HIV care clinics, with high ART and care drop-out rates, resulting in poor virological control and high HIV-related mortality. However, to our knowledge, there has not been any prior research to elicit the factors that influence adherence to ART in the CNB. Participants reported several individual/social facilitators and individual/social and structural barriers to ART adherence and retention. These facilitators and barriers in care are based within the interaction of individual/social/structural levels, as described in Bronfenbrenner’s mesosystem (17).

Individual-level factors were based on psychological health, which was mainly related to social acceptance/discrimination. The association of good mental health with social support at the foundation has been described as the backbone of ART adherence (18) and has been shown to impact interpersonal, psychological and mental health, especially anxiety and depression (19, 20).

We found that not all social exclusion affects the participant’s adherence and retention the same. For example, family-level discrimination was detrimental to psychological wellbeing, and therefore to adherence and retention. However, although community-level discrimination affected participants, simultaneous family support seemed to counteract adverse adherence/retention outcomes. Studies in Thailand and the United States reported the importance of family support in ART retention and adherence, especially for young people (21, 22).

Structurally, we document regular ART shortages in the clinics which deter adherence to ART. Additionally, the existence of two medical systems, one Western the other Traditional, that often do not complement each other but instead often compete for patients. Ngäbe-Buglé individuals often chose the geographically and culturally closer provider, albeit more expensive Traditional providers, and attend Western hospitals when the Traditional medicine fails. The interaction between the two medical systems was detrimental to ART adherence and retention in Western HIV care for some participants. Medical pluralism, the use of multiple medical systems such as Traditional and Western systems together or interchangeably, is especially popular in chronic conditions such as HIV (23). Traditional medicine, widespread across the CNB (24), may be favored by patients because of the focus on comprehensive and culturally congruent care (25). Despite existing plurality, patients reported that both systems asked them not to use simultaneous treatments.

### Implications for policy and practice

Families are often overlooked in ART care interventions. However, family systems are central to social functioning in the CNB (26). We found that family support was vital for adherence and retention; interventions with families have been found to support positive HIV outcomes (21, 22). Broader social-network and community-level approaches could increase societal knowledge and support while decreasing stigma, as shown in Kenya and Zimbabwe (27, 28). These interventions could be part of existing HIV care in order not to lose patient continuity. In CNB, some participants mentioned the success of the professionally guided peer-support group; this group should be revived and expanded. In Zimbabwe, the Friendship Bench, a low-cost therapy intervention run by lay healthcare workers, was found to be effective mental-health support for PLHIV (29); in CNB, a similar program could incorporate individuals and family members.

A major structural limitation related to ART access was influenced by cost and distance to travel to the clinic. This limitation was, without a doubt, exasperated by the COVID-19 pandemic in 2020 and 2021, where the *Covidization* of healthcare significantly influenced HIV care (30). We found that patients were expected to retrieve ART monthly and participate in biannual CD4 count/HIV plasma viral load check-ups during our data collection. Although ART in Panama is free (5), monthly clinic trips may be too much of a financial burden to continue treatment, as found in a worldwide review (31). Home-based visits in China, Uganda, and Rwanda have increased adherence and viral suppression (32-35). Task shifting to dispense ART through non-specialized health workers could decrease travel and increase adherence and clinical outcomes, as shown in several studies (35-39). However, newer research has found community-based drug delivery could be costly to the clinics and could force clients to identify as living with HIV (40). Other models of differentiated delivery of ART include fast-track drug refills for 3-6 months. In the CNB, fast-track, multi-month refills and decentralized services to smaller health centers across the country could decrease the time and money spent on monthly pill pick-up.

Food insecurity has been shown in a worldwide review as a limiting factor to ART adherence due to more significant side effects of ART when taken without food and competing for economical use demands when choosing between clinic travel costs or food (41). Despite existing supplemental nutrition in CNB, some still felt food scarcity. Therefore, the provision of other food items should be considered. A study in Honduras found an increase in adherence when a monthly food basket was given at ART retrieval (35, 42).

In CNB, the relationship between Western/Traditional medicine was found to be exclusionary. However, these systems could coexist for successful patient care, as found in South Africa (43). Successful collaborations could decrease potential morbidity and mortality related to potentially unsafe practices and low adherence/retention (43-45). Further complicating these interactions is the promise of cure by some Traditional providers in CNB; similar findings have emerged in Uganda (46) and Tanzania (47). Due to the overwhelming use of Traditional medicine throughout the CNB, despite the high costs, Western medicine could benefit from the trust and cultural closeness by systematically including Traditional providers to help deliver care. Interventions could incorporate Traditional providers for ART promotion and patient home visits. A study in Malawi found that adherence and retention increased with the inclusion of Traditional providers as community health workers (48). However, in Zambia, despite Western and Traditional providers recognition of the importance of collaboration, institution-based, one-way teaching methods were unsuccessful in promoting collaboration (49).

In this study, we found consistent findings with a diverse group of PLHIV, including adherent/somewhat non-adherent patients. We also included a nearly identical participant sex ratio to CNB clinic statistics, as 82% of PLHIV in CNB are male (50). Additionally, code saturation was reached quickly. However, this study has two main limitations, firstly the interviewers, AG and ES, are ethnically white women, which may have decreased accurate reporting among Indigenous participants. However, HIV status in CNB is generally seen as a sensitive and confidential topic; the fact that the interviewers were markedly from outside the CNB may have increased openness as the risk of divulging serostatus among community members decreased. Secondly, ART clinicians aided in the selection of participants; selection bias may have occurred. Furthermore, we could not contact completely non-adherent participants. However, participants presented with a mixture of adherence levels from adherent to somewhat non-adherent.

## Conclusions

This qualitative study among PLHIV focused on a relatively isolated group of Indigenous peoples who live in the CNB in Western Panama. We found that participants’ experience in ART adherence and retention in care was strongly influenced by interrelated individual, social, and structural factors. These factors included individual psychological health as well as family and community social support or discrimination. Notably, PLHIV in the CNB have difficulties in deciding between Traditional and Western medicine offers because of distance of travel and cultural closeness. Additionally, poverty and food insecurity made adherence difficult for many, while others were affected by regular ART shortages. We provided several recommendations from other studies in Low- and Middle-income countries, including interventions that impact the social and structural-level barriers to adherence and retention in ART care.

## Data Availability

All data produced in the present study are available upon reasonable request to the authors

## Acknowledgments

We are incredibly grateful to the participants who took the time to meet and talk to us about their lives. We are also grateful to the Ministry of Health and Traditional Doctors of the Comarca Ngäbe-Buglé for supporting and encouraging this study. In addition, JMP is a distinguished member of the SNI of SENACYT.

## Co-author Contributorship

| **Contributor Role** | **Role Definition** |
| --- | --- |
| **Conceptualization** | AG, ES, CG, YQ |
| **Data Curation** | AG, ES |
| **Formal Analysis** | AG, ES |
| **Funding Acquisition** | AG, JMP |
| **Investigation** | AG, GCT, AC, YQ, CG |
| **Methodology** | AG, PM |
| **Project Administration** | AG |
| **Resources** | GCT, AC, YQ, CG |
| **Software** | N/A |
| **Supervision** | JMP, PM |
| **Validation** | PM |
| **Visualization** | AG, PM |
| **Writing – Original Draft Preparation** | AG, ES, JMP, GCT, AC, YQ, CG, PM |
| **Writing – Review & Editing** | AG, ES, JMP, GCT, AC, YQ, CG, PM |

